# A standard scoring method for measuring white cast of mineral sunscreens and improving user compliance across diverse skin tones

**DOI:** 10.1101/2025.02.12.25322144

**Authors:** Alexandra M. Maldonado López, Emily Gallagher, Aiden Curry, Kylie Q. Sahloff, Ivan Domicio da Silva Souza

**Affiliations:** Good Molecules, LLC, Philadelphia, Pennsylvania, United States of America

## Abstract

Using broad-spectrum sunscreen is an effective practice for preventing skin cancers caused by ultraviolet (UV) radiation. Mineral sunscreens containing zinc oxide (ZnO) and titanium dioxide (TiO_2_) as physical UV filters are suitable for individuals with sensitive skin or allergies to chemical UV filters. Consumer compliance with sunscreen application depends largely on its cosmetic elegance, especially regarding white cast. Despite this, there is no official method to quantify white cast and help design sunscreens for diverse skin tones. To address this gap, we developed validated white cast scoring protocols that combine 1) subjective consumer feedback on formulations containing ZnO concentrations ranging from 0–30% with 2) objective L* measurements (whiteness) determined after sunscreen application. Our findings demonstrate a strong correlation between increasing ZnO percentages and higher L* values, resulting in a more visible white cast. White cast scores for the ZnO formulations were substantially consistent across both in vivo and in vitro methodologies, with higher ZnO concentrations producing unacceptable levels of white cast. This study provides a white cast scoring system as a quantitative tool for evaluating and refining mineral sunscreen formulations, contributing to the development of more cosmetically elegant sunscreens suitable for a wide range of skin tones.

## Introduction

Ultraviolet radiation (UVR) is the primary driver of several skin cancers [1]. Recent estimates state that one in five Americans will develop skin cancer in their lifetime [2]. Broad-spectrum sunscreen usage is an effective measure for preventing skin cancer [3]. Daily application of broad-spectrum sunscreen protects the skin against ultraviolet A (UVA) and ultraviolet B (UVB) when exposed to UVR [1]. Sunscreens are divided into two categories based on the presence of their UVR filter actives: chemical (also known as organic) filters or physical (also known as inorganic but commonly known as mineral) filters. Mineral sunscreens, which consist of the two U.S. Food and Drug Administration (FDA) approved physical filters, zinc oxide (ZnO) and titanium dioxide (TiO_2_), are safe and effective options for sun protection. ZnO and TiO_2_ are physical filters that scatter, reflect, and absorb UVR [4]. Some state that TiO_2_ effectively absorbs primarily UVB, while ZnO mainly absorbs UVA radiation, whereas combining both provides broad UVA/UVB protection [5]. Notably, at higher concentrations (> 7%), ZnO alone can provide broad-spectrum protection, as well as protection against long-wavelength UVA [6]. ZnO is a gentle option for sunscreens, making it suitable for individuals with sensitive skin or allergies to UV chemical filters.

One of the most desired properties consumers seek in sunscreen is its cosmetic elegance for an aesthetically pleasing look (the way sunscreen has easy spreadability and is invisible when applied to the skin). However, consumers tend to stray away from mineral sunscreens due to their lack of cosmetic elegance since ZnO and TiO_2_ leave a white residue on the skin, called white cast [7]. White cast from mineral sunscreens often deters consumers, especially those with darker complexions [8,9]. Dermatologists recommend applying sunscreen at 2 mg/cm^2^ every two hours to protect against the harmful UVR [9]. This amount of sunscreen needed for protection can cause a substantial white cast, leading consumers to apply sunscreen incorrectly or even avoid application, exposing them to higher skin cancer risk. As a result, when consumers are counseled on sunscreen use, most of the recommended sunscreens contain chemical filters that are transparent when applied [10,11]. On one hand, UV chemical filters were found to be harmful to aquatic environments, causing coral bleaching and marine life toxicity [12–15]. Environmental impacts have led to the banning of chemical filters in several territories to protect islands and coastal environments [16]. On the other hand, dermatologists have expressed concerns that ingredient restrictions might lead people to use sunscreens less frequently [16]. Understanding and addressing these consumer and environmental concerns is essential to ensure satisfactory and consistent use of sunscreens. For this reason, industries are designing cosmetically elegant “reef- safe” sunscreens based on mineral ingredients such as ZnO [17].

ZnO and TiO_2_ have been used in sunscreens since at least the 1940s [18], and their development has been scrutinized over the years. ZnO and TiO_2_ particles used in sunscreens form aggregates that can agglomerate between 0.1–10.0 μm in size [19] through their innate chemical and physical properties. The larger the agglomerate, the more direct impact on UVR absorption and the resulting sun protection factor (SPF) value [20]. Also, the bigger the agglomerate, the more obvious the white cast is [21] since more particles reflect and scatter wavelengths in the visible light range, which the retina perceives as white [4]. Thus, since ZnO and TiO_2_ particles form aggregates and agglomerates in sunscreen formulations, it is known that increasing concentrations of ZnO and TiO_2_ particles in sunscreen formulations can lead to increased white cast [7]. ZnO and TiO_2_ aggregates clump together primarily during manufacturing, where aggregates are exposed to heat and drying processes, forming the agglomerates, which result in a sunscreen that reflects more visible light and leaves a white cast on the consumer [22]. A way to deagglomerate metal oxide particles is via milling, ultrasonication, or functionalization, which leads to repulsion or reduced attraction between agglomerates. Interventions in the formulation stage that directly correlate with particle size and interparticle interactions can be done to reduce white cast. An example is to implement nanoscale metal oxides [7]. Additionally, pre-prepared ZnO and TiO_2_ dispersions have been used to inhibit the formation of larger aggregates and agglomerates typically produced with free-flowing powders [7,23].

Even though there is no official method to measure white cast objectively, quantitative transparency measurements can be used to optimize formulation development. An example is determining the size-dependent transparency of the metal oxides through the Lorenz–Mie–Debye theory. This theory has been used to plot transparency versus the average particle diameter of TiO_2_ to predict the aesthetic appeal of sunscreens that use nanoparticles in vitro [24]. Other work has also been done to quantify white cast through a consistent and reproducible skin color evaluation [25,26], which has been helpful for dermatology and cosmetic science research. This skin color assessment in various skin types can be achieved with noninvasive devices through colorimetry by analyzing the intensity of the reflected wavelength to deduce the color it is “seeing”. Colorimeters are an objective color quantification tool that represents human color vision developed under the standardization of the Commission Internationale de l’Eclairage (CIE), an international authority on light and color [27]. Their color quantification can be represented under many color systems. The 1976 CIEL*a*b* color system is widely used for more in-depth research. It operates under the premise of opponent-process theory with a three-dimensional color space. The L* axis is grayscale with 0 (black) to 100 (white) values, which correlates with an individual’s level of pigmentation. The a* is the red-green axis that correlates with erythema because a positive a* describes red, while a negative a* describes green values. The b* correlates with pigmentation and tanning. It is the yellow-blue axis, in which a positive b* is yellow and a negative b* value is blue [28,29].

Relying on colorimetry to objectively measure white cast can be challenging to reproduce because of the variability in the size distribution, particle shape, and density of the metal oxide particles in sunscreens when spread on the skin, as well as the fact that white cast is perceived subjectively differently across various skin tones. Because of the lack of practical and validated methods for measuring white cast caused by mineral sunscreens in different skin tones, we have created a procedure for determining and recording the white cast of mineral sunscreens containing zinc oxide via CIEL*a*b* in vivo and in vitro, considering the volunteer’s perception of white cast. These methods are intended to optimize the development of mineral sunscreens by allowing the selection of formulations with a reduced white cast at the early stages of the product development cycle.

## Results

### Model fit and method validation

In vivo and in vitro experiments are essential when formulating mineral sunscreens. Clinical studies evaluate the white cast produced by mineral sunscreens across different skin tones before the product reaches the market. Meanwhile, in vitro testing offers a cost-effective method for screening and selecting promising sunscreen formulations before clinical trials. Thus, we wanted to establish protocols to reliably measure white cast in vivo and in vitro. The white cast measurements in our protocols focus on the L* value of the CIEL*a*b* color system since its linear values span from black to white. For the in vivo protocol, we compared test formulations containing a range of ZnO (0-30%) on volunteers of different Individual Typology Angle (ITA°) subtypes (S1 Fig). When relevant, we further divided the volunteers’ ITA° subtypes into three skin pigmentation categories: **light skin pigmentation** (n = 5, comprising Very Light and Light ITA° subtypes), **medium skin pigmentation** (n = 3, comprising Intermediate and Tan ITA° subtypes), and **dark skin pigmentation** (n = 5, the Brown ITA° subtype only). For the in vitro protocol, we employed VITRO-SKIN over a dark or light background as the substrate for the sunscreen application. The benchmark (BM) of 14.7% ZnO was not included in the model fit and validation to prevent compounding effects due to being formulated differently than the 0-30% ZnO formulations, such as having different sizes or shapes of the ZnO particles.

We performed linear regression analysis to investigate if the ZnO percentage explains the L* variance in the test formulations by assessing the goodness of fit in vivo and in vitro. For the analysis in vivo (Fig 1A-1D), when the measurements from all volunteers are grouped, the adjusted R^2^ of 0.261 indicates a poor fit, suggesting that the model explains only a portion of the variance in L* values based on ZnO percentage. The model’s performance improves when the data is analyzed separately by skin pigmentation categories. The fit was moderate in volunteers with light skin pigmentation, with ZnO percentage accounting for some of the variance in L* values. The model showed a good fit for medium skin pigmentation, effectively capturing most of the variance in L* and performing reliably. In the dark skin pigmentation category, the fit was particularly strong, with the model explaining the majority of the variance in L*, reflecting a robust and highly effective relationship between ZnO percentage and L*. Furthermore, the measurements on VITRO-SKIN with a black background resulted in an adjusted R^2^ of 0.722, reflecting a good fit between ZnO percentage and L* values (Fig 1E). The VITRO-SKIN on a white background had similar results (S2 Fig).

**Fig 1.**
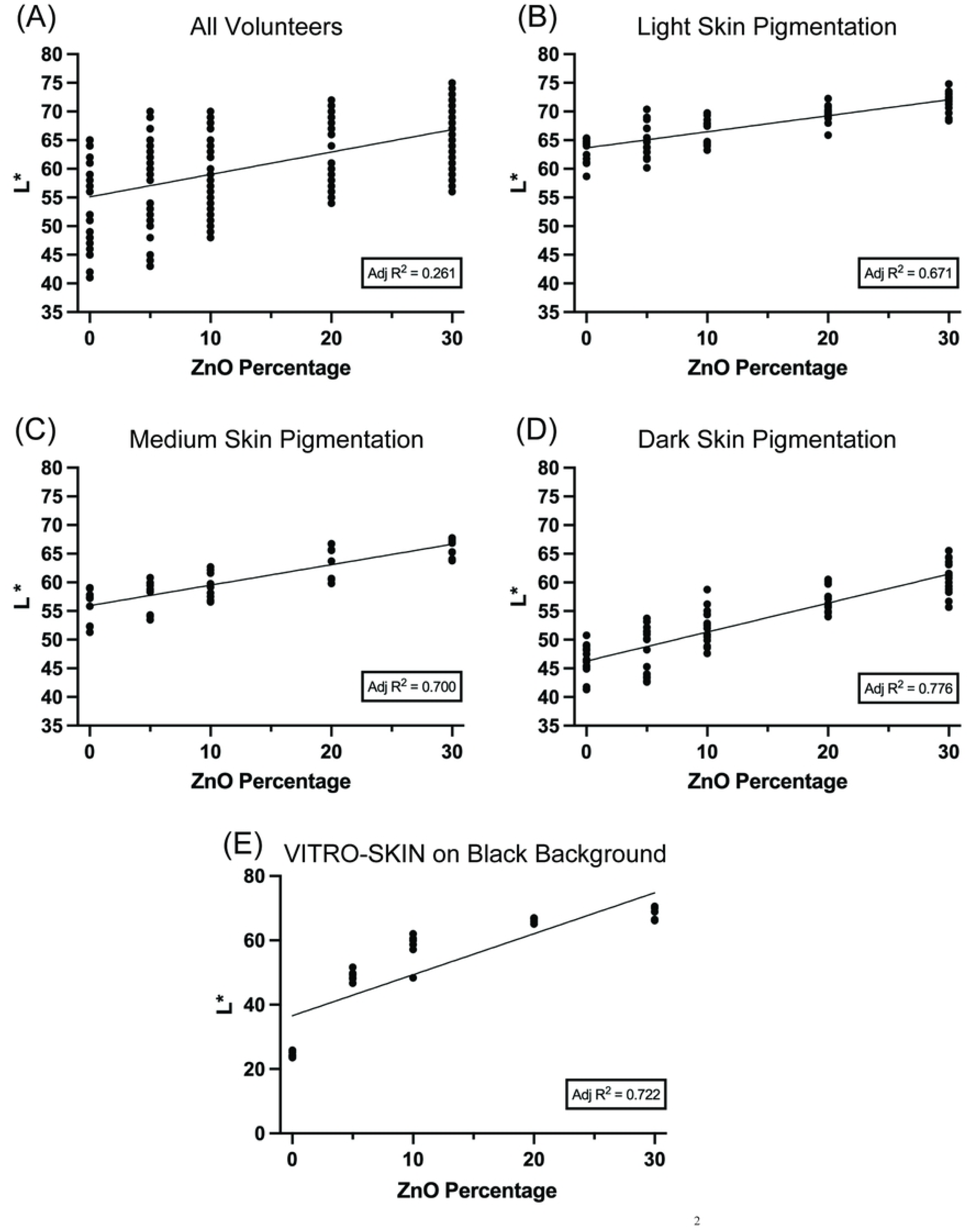
L* value Increases with Increasing Zinc Oxide Percentage. Adjusted R^2^ was calculated to indicate the goodness of fit. (A) All Volunteers’ L* value measurements (n = 195) extend from 41 to 75. (B) Light skin pigmentation volunteers’ L* measurements (n = 75) range from 59 to 75. (C) The L* value measurements of medium skin pigmentation volunteers (n = 45) span from 51 to 68. (D) Volunteers with dark skin pigmentation have L* value measurements (n = 75) from 41 to 66. (E) VITRO-SKIN on black background L* measurements range from 24 to 71 (n = 30).

To continue our validation, a Shapiro-Wilk test was conducted to evaluate the data distribution before performing a correlation analysis between the L* values and ZnO percentages. Since our in vivo data was normally distributed, we performed Pearson correlations to determine the strength of the correlation between ZnO percentage and L* (Table 1). Interestingly, when the volunteers’ measurements were evaluated altogether, there was a moderate correlation between ZnO percentage and L* value, r = 0.520 (p < 0.001). Then, we calculated the Pearson correlation for the three skin pigmentation categories. Light, medium, and dark skin pigmentation volunteers had an r ≥ 0.822 (p <0.001), indicating strong correlations between ZnO percentage and L* when analyzed per skin pigmentation group. An increase in correlation (r-value) was observed with higher skin pigmentation (a darker background for the white cast). For the in vitro measurements, because they were not normally distributed, we performed a Spearman rho analysis (Table 1), resulting in a strong correlation between the ZnO percentage and the L* values, ☐ ≥ 0.938 (p < 0.001).

**Table 1.** Model Performance and Linearity Metrics In Vivo and In Vitro.

| Parameter | All Volunteers | Light Skin Pigmentation | Medium Skin Pigmentation | Dark Skin Pigmentation | Black Acrylic | White Acrylic |
| --- | --- | --- | --- | --- | --- | --- |
| Linearity Range (ZnO %) | 0 - 30 | 0 - 30 | 0 - 30 | 0 - 30 | 0 - 30 | 0 - 30 |
| Regression Equation | $y = 0.3910x + 55.11$ | $y = 0.28x + 63.7$ | $y = 0.357x + 55.9$ | $y = 0.506x + 46.3$ | $y = 1.27x + 36.6$ | $y = 0.0922x + 87.2$ |
| Slope ± SD | $0.3910 \pm 0.04625$ | $0.2801 \pm 0.02270$ | $0.3570 \pm 0.03506$ | $0.5057 \pm 0.03311$ | $1.274 \pm 0.1458$ | $0.09224 \pm 0.008862$ |
| Intercept ± SD (L*) | $55.11 \pm 0.7808$ | $63.66 \pm 0.3832$ | $55.94 \pm 0.5918$ | $46.28 \pm 0.5550$ | $36.59 \pm 2.461$ | $87.22 \pm 0.1496$ |
| Pearson ($p < 0.001$) | 0.520 | 0.822 | 0.841 | 0.883 | | |
| Spearman rho ($p < 0.001$) | | | | | 0.938 | 0.951 |
| n | 195 | 75 | 45 | 75 | 30 | 30 |

We wanted to ensure that the measurements made by the NYX Pro 2 Color Sensor colorimeter were accurate and precise. In vivo, accuracy was calculated by comparing the L* values of all ITA° subtypes present in the study (Very Light to Brown subtypes) to the mean values of L* for ITA° subtypes previously published [25]. All accuracy (Table 2) was between 88-96% of the reference values. Furthermore, precision calculations were made for all ITA° subtypes and the three subsets of Light, Medium, and Dark skin pigmentation (Table 3 and S1 Table). The Relative Standard Deviations (RSDs) of the L* before and after the sample application were < 10%. In vitro, accuracy was analyzed by measuring CIEL*a*b* of pure black and white standard paints on VITRO-SKIN with a black background in comparison with CIEL*a*b* measurements of theoretical pure black (L* = 0, a* = 0, b* = 0*) and theoretical pure white (L* = 100, a* = 0, b* = 0). The accuracy of measuring black on a black acrylic background was 89%, while for white, it was 93% (Table 2). We also calculated the inter- and intra-day precisions (Table 3 and Supplemental Table 2) of our L* measurements on VITRO-SKIN with black acrylic as the background, resulting in RSDs < 7% before and after the test formulation application. The same measurements were taken and analyzed for VITRO-SKIN on a white background, yielding similar results (Tables 2 and 3 and S2 Table).

**Table 2.**
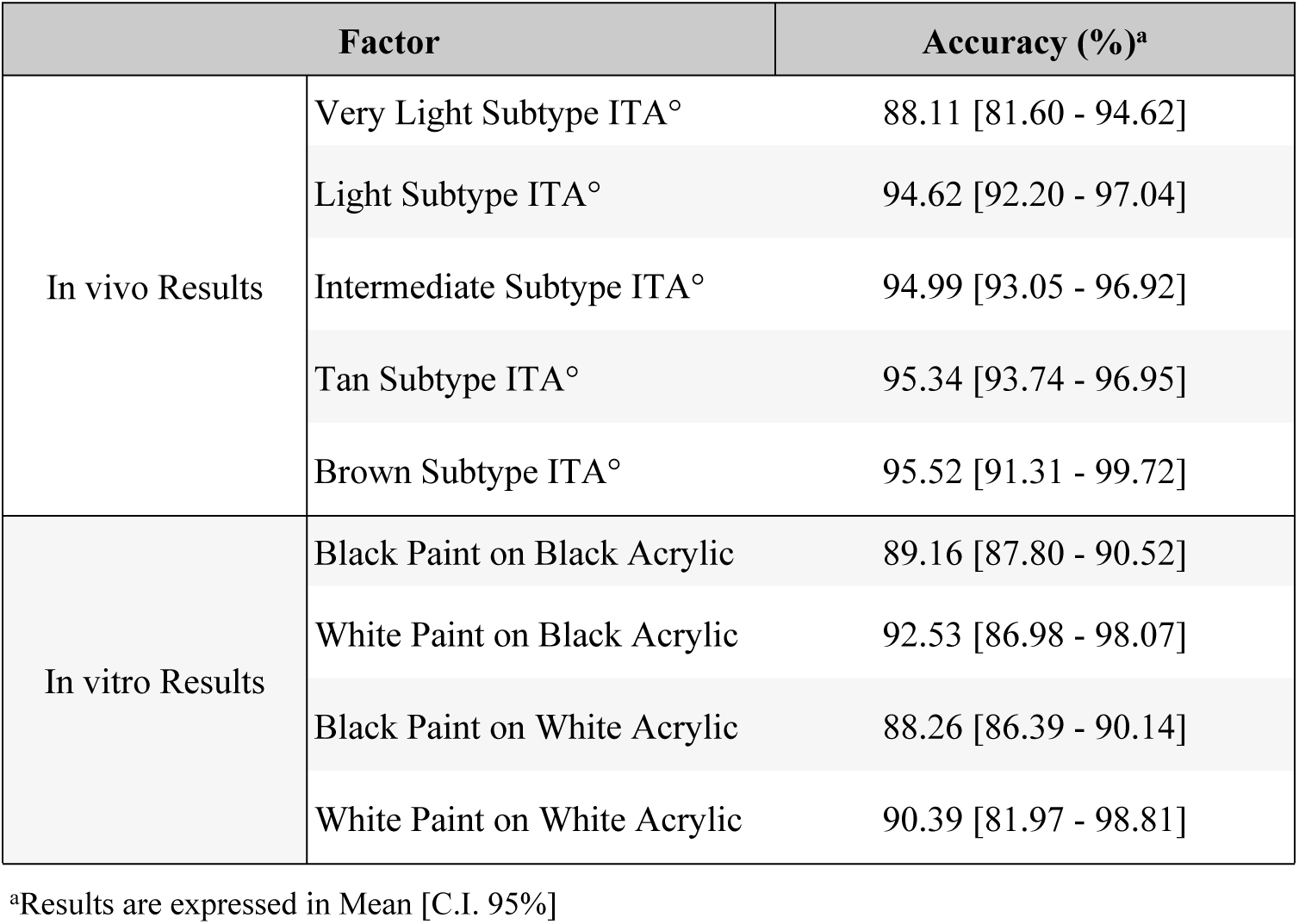
Accuracy of L* Measurements for the In Vivo and In Vitro Protocols.

**Table 3.**
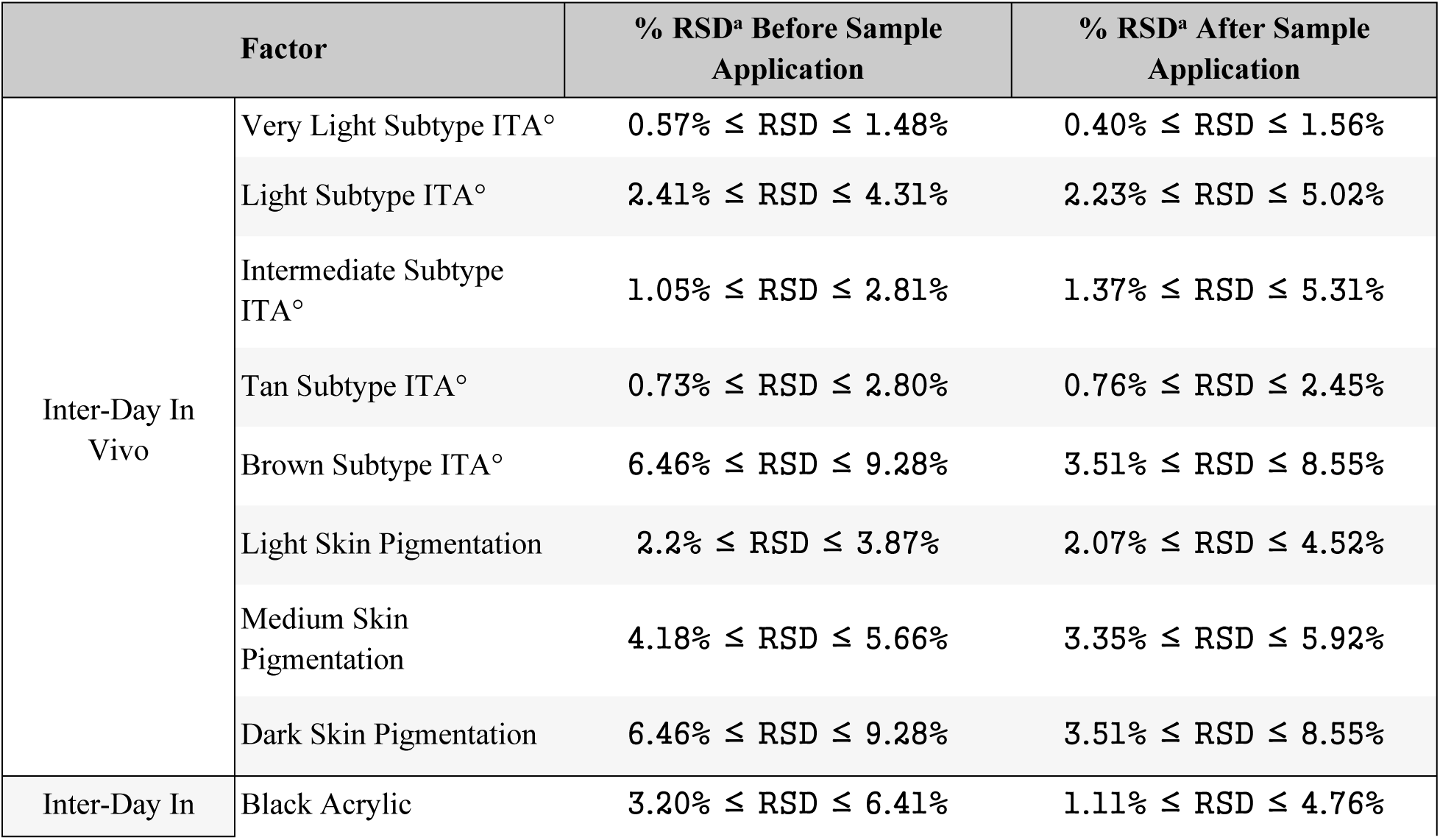
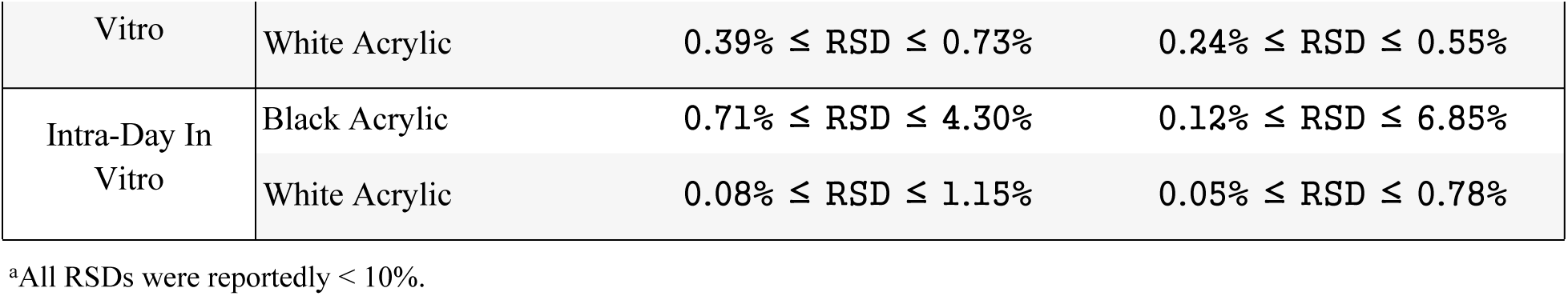
Precision of L* Measurements for the In Vivo and In Vitro Protocols.

### L* measurements

The in vivo study was carried out with a panel of thirteen (13) female volunteers, aged 20- 60 years, Fitzpatrick II-VI, with diverse ethnicities (including 31% Caucasians and 38% Black or African Americans, S3 Table). We recruited volunteers based on their Fitzpatrick Skin Type but analyzed the research based on the volunteers’ ITA° subtypes (Sl Fig). Photographs and CIEL*a*b* measurements were taken of the volunteer’s skin before and after each test formulation application to observe how the skin color changed subjectively (Fig 2). To understand the color variation, we converted the mean CIEL*a*b* measurements into Hexadecimal color codes (HEX) for more convenient observation. In the representative pictures of each ITA° subtype (Fig 2B), we noticed that in the Very Light, Light, and Intermediate ITA° subtypes, the 30% ZnO sample exhibits a prominent white cast on their skin, while, in comparison, the 20% ZnO caused less white cast. In the Tan and Brown ITA° subtypes, the 20% and 30% ZnO samples caused a more apparent white cast, but the 10% ZnO also produced some white cast on their skin tones (Fig 2B). Observing the HEX color values (Fig 2C) shows a grey gradient with the increase in ZnO percentage. The grey gradient can be observed in the Very Light, Light, and Intermediate ITA° with the 20% and 30% ZnO samples, while it is more striking in the Tan and Brown ITA° subtypes, starting at the 10% ZnO (Fig 2C).

**Fig 2.**
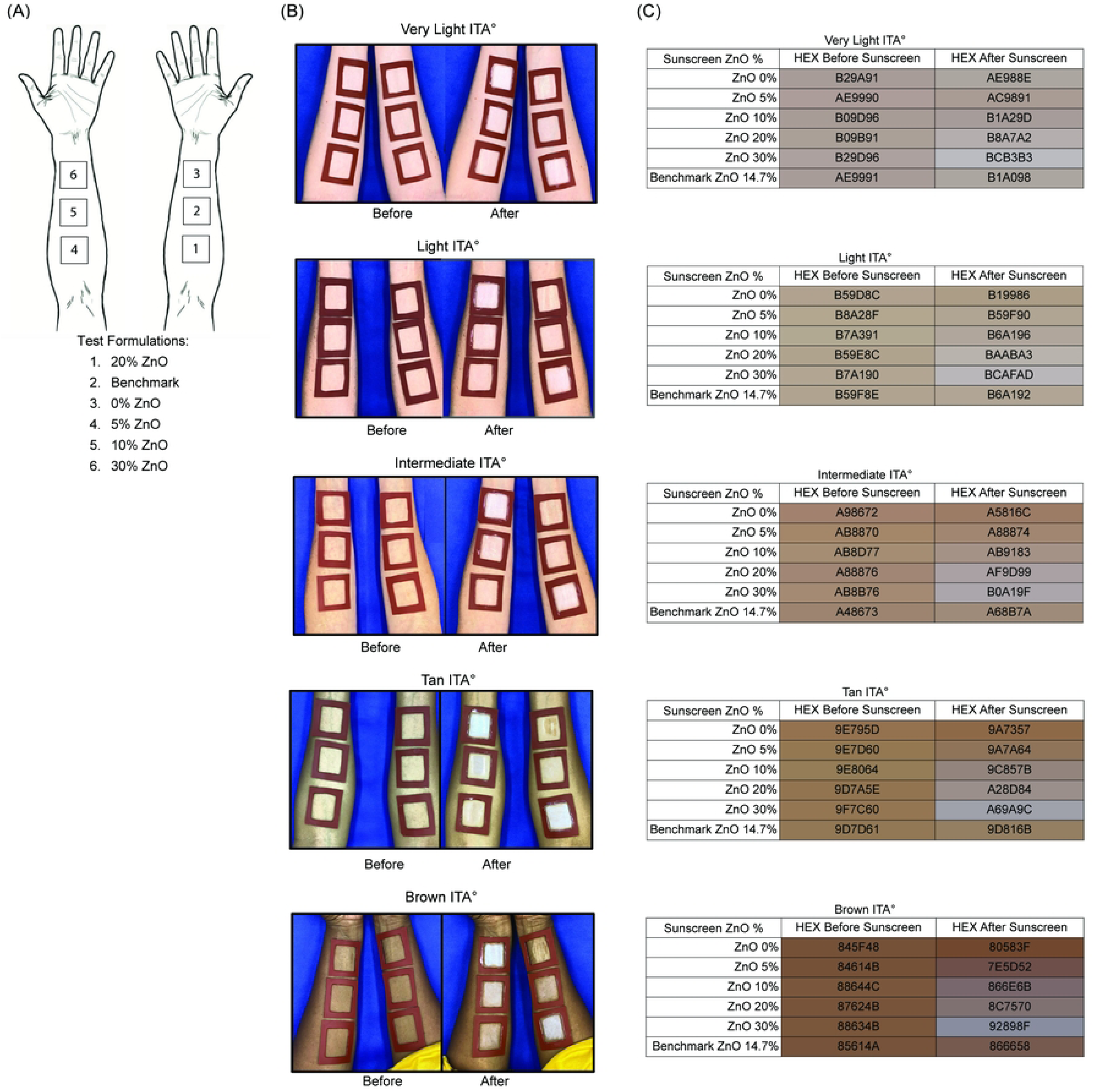
White cast Visualization caused by Different Zinc Oxide Percentages in a Variety of Skin Tones. (A) Legend indicating which formulation was applied to each square on the volunteers’ forearms. (B) Representative images of how white cast caused by different zinc oxide percentages are experienced by each ITA° subtype. (C) Average HEX skin color of each ITA° subtype before and after the application of each test sample.

### Subjective white cast perception

Apart from subjectively visualizing the white cast difference caused by each test formulation, we quantified the whiteness change caused by each ZnO percentage (Fig 3). One-way ANOVA was performed to determine the objective shift in whiteness (L* value) caused by ZnO percentage before and after the test formulation application. All volunteers had a significantly different increase in L* value after applying the 20% and 30% ZnO sunscreen formulations (p < 0.001 and < 0.0001, respectively, Fig 3A). Since we observed subjectively that the formulations do not cause the same white cast in different skin tones and the baseline of L* of each subcategory spans different L* values [25], we decided to continue our analysis on the three skin pigmentation categories: light, medium and dark. When this subdivision was analyzed, we noticed that the 20% ZnO difference was more statistically significant (p < 0.0001) in all three skin pigmentation categories, while the 30% ZnO maintained the same p-value (Figs 3B-D). Similar to the in vivo study, we quantified the whiteness shift caused by each ZnO percentage on VITRO-SKIN with a black acrylic sheet as the background (Fig 3E). A statistically significant difference between L* before and after the formulation applications was observed for 5-30% ZnO percentages, including the BM, with a significance of < 0.0001.

**Fig 3.**
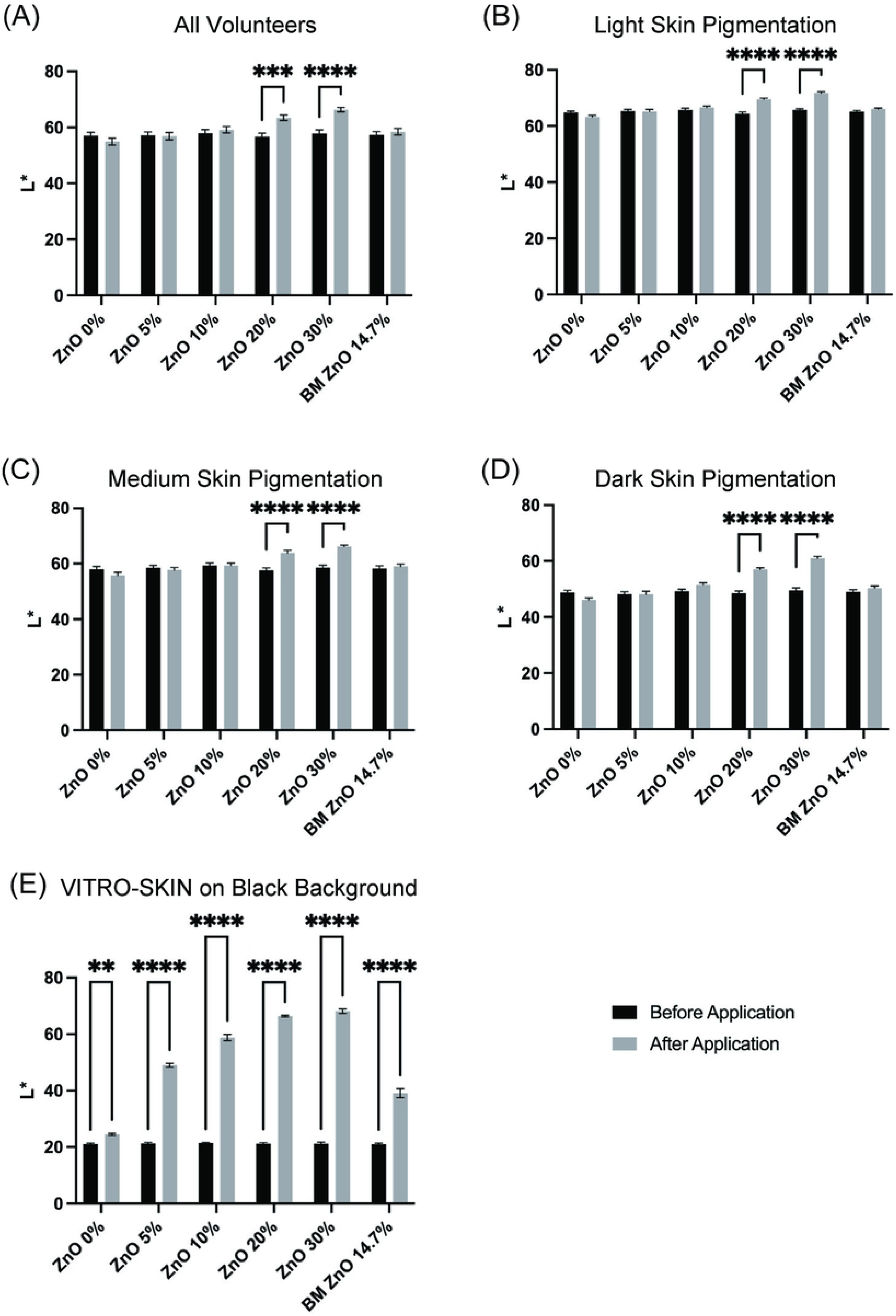
L* value Increases after Application of Zinc Oxide Formulations. In vivo: (A) 20% and 30% ZnO test formulations show statistically significant changes in L* values after their use in All Volunteers (p < 0.001 and p < 0.0001, respectively, n = 39). (B-D) Light skin (n = 15), medium skin (n = 9) and dark skin pigmentation (n = 15) volunteers show statistically significant changes in L* value when using 20% and 30% ZnO test formulations (p < 0.0001). (E) In vitro: 0% ZnO test formulation has a statistically significant difference after its application (p < 0.01). Test formulations with ZnO percentages ranging from 5-30% and the 14.7% benchmark (BM) show statistically significant changes in L* values after their application (p < 0.0001). n = 6. Error bars are shown as mean ± SEM.

A subjective questionnaire was administered to gauge the volunteers’ opinions on white cast, resulting in 85% expressing concerns about white cast on their skin when applying sunscreen. Questions about the test formulations were also administered (Table 4). The volunteers ranked the formulations on their forearms from least to most white cast. An Interclass Correlation Coefficient (ICC) was calculated for their ranking to explore if there was consistency in how the volunteers perceived white cast on their skin. Volunteers had difficulty discerning the white cast between the 5% ZnO and BM 14.7% ZnO formulations. Nonetheless, we obtained an ICC of 0.991 (p < 0.001) with a 95% confidence interval between 0.975 and 0.999, indicating a remarkably high level of ranking agreement amongst volunteers. This high level of agreement provides a foundation for the subsequent questions. When asked about the maximum level of white cast they would accept on their face in exchange for sun protection, 23% of volunteers preferred a sunscreen with 0% ZnO, 38% were comfortable with 5% ZnO or the BM sunscreen, 31% preferred 10% ZnO and only one volunteer was willing to accept a 30% ZnO white cast. The preferences shifted when the same question was asked regarding white cast on the body. Only one volunteer preferred 0% ZnO, 31% were comfortable with 5% ZnO or the BM, 38% preferred 10% ZnO, and 23% were open to sunscreens with a 20% to 30% ZnO level of white cast.

**Table 4.**
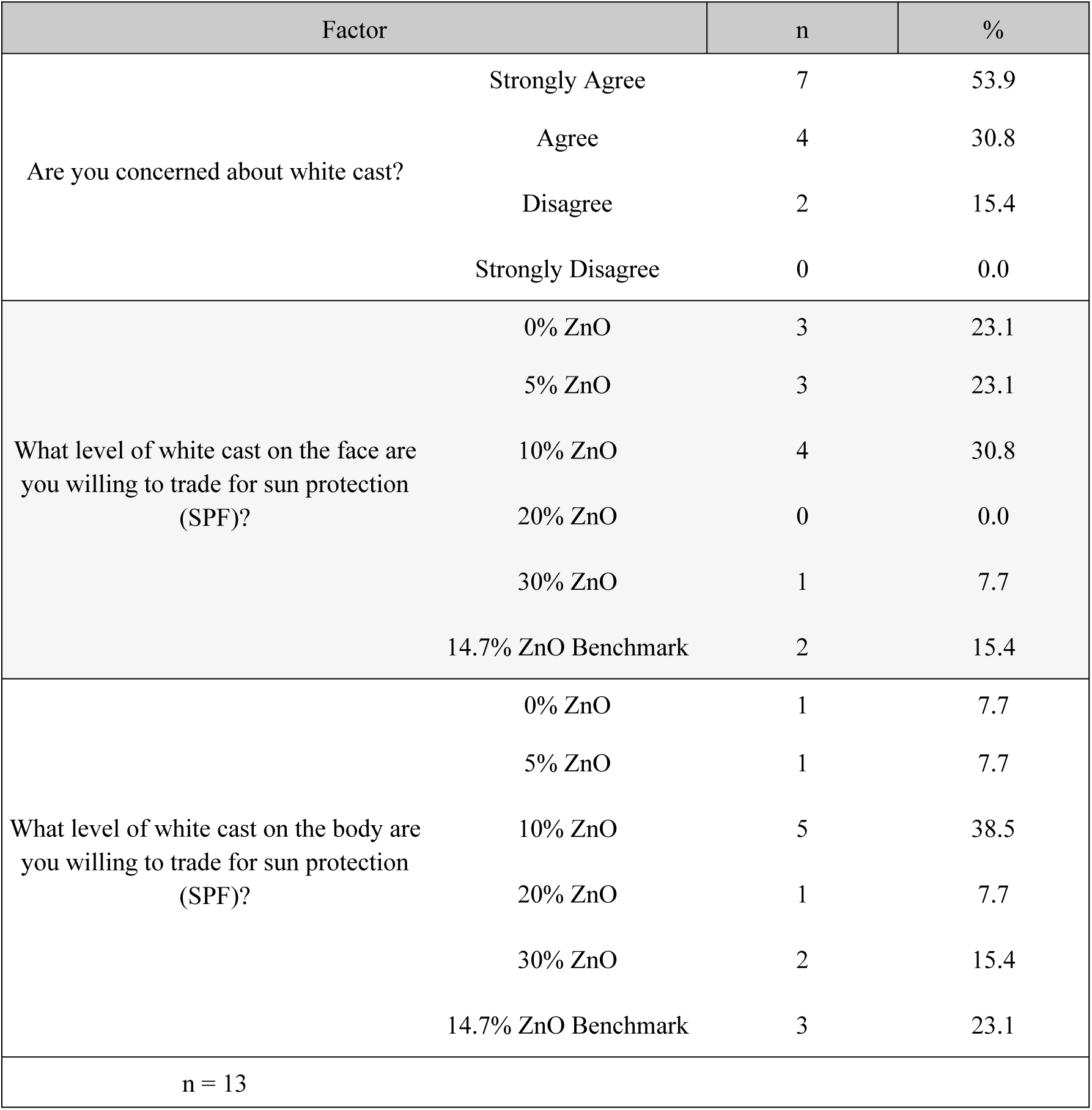
White Cast Subjective Assessment.

### White cast score

There is currently no official method to quantify the amount of white cast caused by sunscreens. To address this gap in sunscreen formulation, we developed a white cast score specifically for mineral sunscreens. The white cast score for each test formulation was calculated according to the equation: 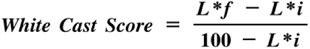 where *L*f* = L* measurement after test formulation application, and *L*i* = L* measurement before test formulation application. The white cast scores for each test formulation and protocol are presented in a graph along with their interpretation (Fig 4).

**Fig 4.**
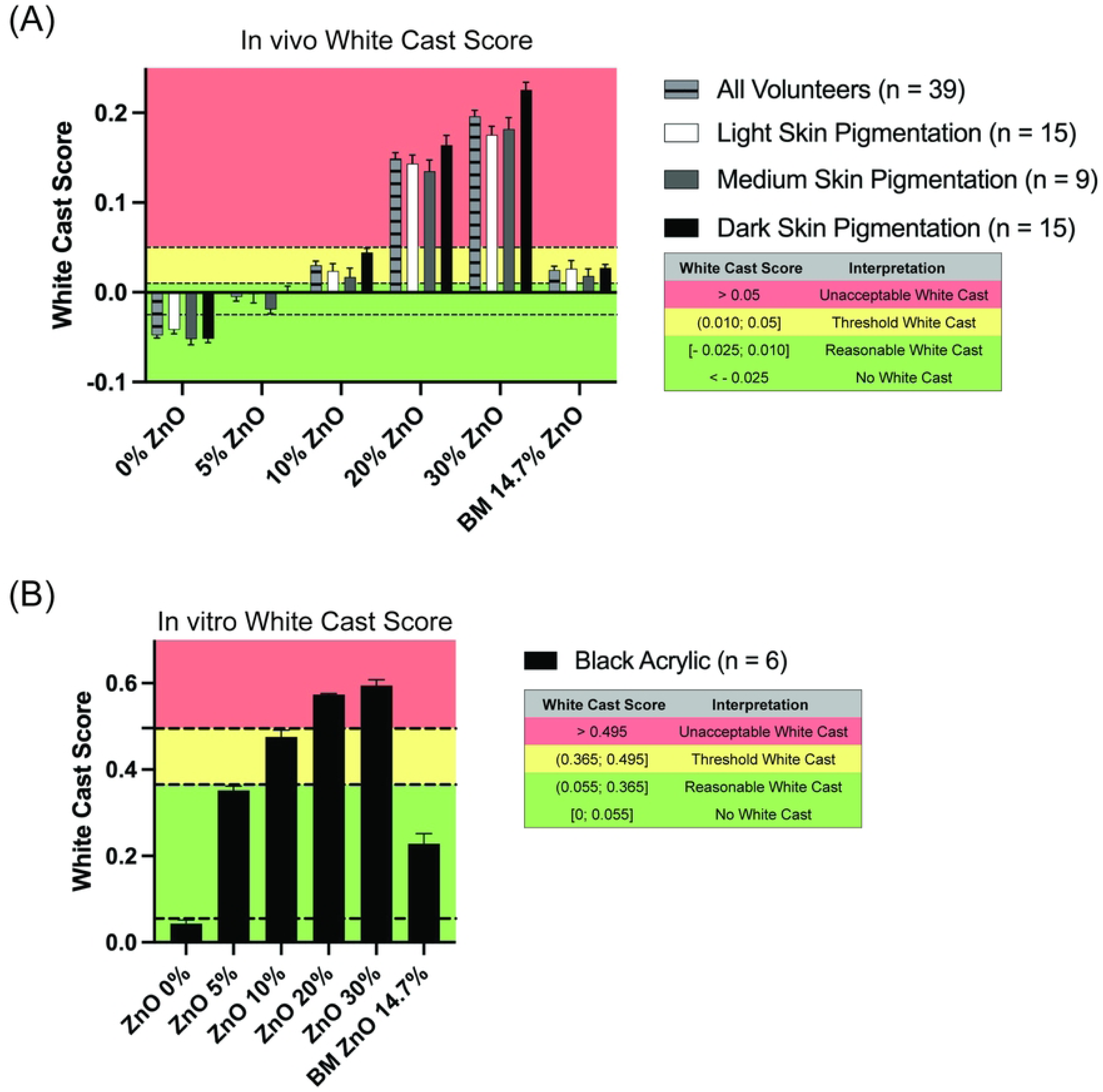
In vivo and in vitro White cast Scores. Scores are categorized as “No White Cast”, “Reasonable White Cast”, “Threshold White Cast” and “Unacceptable White Cast” based on a combination of L* values and volunteer feedback. (A) White cast scores determined for in vivo measurements. (B) White cast scores determined for in vitro measurements. BM = benchmark. Error bars are shown as mean ± SEM.

For in vivo (Fig 4A), the negative control test formulation with 0% ZnO showed no white cast on any volunteer. The 5% ZnO test formulation produced a reasonable white cast in all volunteers. The 10% ZnO and the BM (14.7% ZnO) formulations in the majority of skin tones fell into the threshold white cast category, between reasonable and unacceptable white cast levels. Finally, 20% and 30% ZnO formulations received an unacceptable white cast score for all volunteers. For in vitro (Fig 4B), the 0-30% ZnO formulations essentially replicated the in vivo scores. The BM sunscreen differed from the in vivo results by being classified as a reasonable white cast in vitro.

We conducted Pearson correlation tests for in vivo (all volunteers, light, medium, and dark) and in vitro white cast scores (Table 5). Subsequently, we verified whether the correlation coefficients for each in vivo category differed significantly from the in vitro coefficient. Results showed no significant differences between the correlation coefficients for any of the comparisons: All volunteers vs. VITRO-SKIN on black background, Light skin-pigmented volunteers vs. VITRO-SKIN on black background, Medium skin-pigmented volunteers vs. VITRO-SKIN on black background, and Dark skin-pigmented volunteers vs. VITRO-SKIN on black background.

**Table 5.** White Cast Scores for In Vivo and In VItro Protocols Correlate Equivalently.

| Comparison | Pearson Correlations (p < 0.001) | Fisher's Z Conversion | Z difference<sup>a</sup> |
| --- | --- | --- | --- |
| All Volunteers vs. Black Acrylic | 0.904 vs. 0.774 | 1.494 vs. 1.030 | 1.315 |
| Light Skin Pigmentation vs. Black Acrylic | 0.884 vs. 0.774 | 1.394 vs. 1.030 | 0.946 |
| Medium Skin Pigmentation vs. Black Acrylic | 0.919 vs. 0.774 | 1.583 vs. 1.030 | 1.312 |
| Dark Skin Pigmentation vs. Black Acrylic | 0.928 vs. 0.774 | 1.644 vs. 1.030 | 1.520 |

<sup>a</sup>Z differences < 1.96 are not statistically significant at the 0.05% level.

## Discussion

To address the lack of a standardized method for quantifying white cast in sunscreen formulation, we developed a validated protocol to measure the white cast caused by mineral sunscreens both in vivo and in vitro using CIEL*a*b* measurements. In vivo, melanin content in the epidermis correlates negatively with L* values. Therefore, individuals with lighter skin pigmentation have a higher L* value and a higher ITA° color classification than individuals with darker skin tone [27,30–32]. Recognizing this, we divided the volunteers into the light, medium, and dark skin pigmentation categories due to the different baseline L* values for each ITA° subtype. This categorization allowed us to better evaluate the performance of various sunscreen formulations across diverse skin tones.

Formulations used for testing containing 20% and 30% ZnO caused significant white cast visually and quantitatively in light, medium, and dark skin pigmentation categories. Interestingly, the 10% ZnO test formulation exhibited white cast visually in photos of Tan and Brown subtypes despite the absence of statistical significance in the L* shift. This observation positioned the 10% ZnO test formulation within a critical range for white cast quantification. Additionally, the 5% ZnO and BM test formulations are particularly intriguing, as there is no statistically significant difference in the L* shift before and after their application, and regardless of skin tone, volunteers could not visually distinguish between the 5% ZnO and BM formulations. This highlights the complex interrelationship between measurable white cast and consumer perception. Notably, in vitro measurements using VITRO-SKIN with black acrylic backgrounds detected significant L* value changes across all ZnO concentrations, confirming its sensitivity and reliability.

To complement these objective white cast measurements obtained, we administered a subjective questionnaire to volunteers to evaluate consumer preferences and needs regarding the cosmetic elegance of sunscreens. Volunteers prefer formulations with minimal white cast on their faces when choosing sunscreen, while they are more willing to tolerate white cast on their bodies if the sunscreen provides effective sun protection. This difference in the preference for white cast on the face versus the body can be due to aesthetics and cosmetic elegance. A volunteer expressed this sentiment: “It [white cast by mineral sunscreens] does not bother me too bad on the rest of my body, but it is quite obvious on my face”.

These insights, along with the L* value measurements, informed our development of a white cast score with four categories: “no white cast”, “reasonable white cast”, “threshold white cast”, and “unacceptable white cast”. The threshold white cast category is a borderline range that falls between being reasonable (cosmetically acceptable) and unacceptable (not aesthetically pleasing). Sunscreen formulations with a white cast score in this range function like an ambiguous element that may be subjectively perceived as either reasonable or unacceptable, depending on skin tone and individual preferences. Our analysis showed that the 0-30% ZnO test formulations had consistent white cast scores in vivo and in vitro, with the higher ZnO percentages having unacceptable white casts (20-30%). However, 5% ZnO and BM test formulations fall into different scores (reasonable vs threshold white cast, respectively) in vivo, even though volunteers could not visually distinguish between them. This highlights the discrepancy between objective measurements and the consumer’s perception. Interestingly, the in vitro method proved more effective in identifying the BM test formulation as having a reasonable white cast score, aligning more closely with volunteer preferences in white cast on their skin. Moreover, no significant difference between the direction of in vivo and in vitro white cast scores was observed, indicating that the white cast scores for both protocols correlate equivalently. This emphasizes the in vitro method as a reliable tool for predicting consumer perceptions when optimizing sunscreen formulations.

We have developed a validated protocol for measuring and recording the white cast of mineral sunscreens containing ZnO with all in vivo and in vitro L* achieving > 88% accuracy and being satisfactorily precise, with an RSD of < 10%. This white cast measurement protocol can assess white cast in vivo for a finished formulation or in vitro while formulating mineral sunscreens containing ZnO, and potentially other white pigments, such as TiO_2_. Implementing this protocol requires consistent lighting and careful calibration of the colorimeter to ensure accurate results that accommodate diverse skin types and ethnicities. Our study has provided valuable insights; however, it is essential to note its limitations. When measuring the ZnO 0% test formulation with black acrylic as background, statistically significant differences exist between the L* values when comparing the before and after measurements (p < 0.01). This suggests that the other ingredients in the sunscreen formulations, not UV filters, may also affect the L* values. Additionally, we used one ZnO ingredient among the many types available for formulating. Different kinds of ZnO particles produce different white casts due to the manufacturing process altering their shapes, sizes, and transparency. For example, the BM ZnO sunscreen scored as threshold white cast in vivo but as reasonable white cast in vitro. This discrepancy may be due to how the ingredients of the BM formulation interact with the VITRO-SKIN model. Another limitation lies in our volunteer pool, even though the in vivo study proved successful in measuring white cast in a wide range of skin types in the ITA° objective color classification, we could not recruit volunteers with a Dark ITA° subtype. This limitation originates from the historical inconsistency of the Fitzpatrick Skin Type classification, which was originally developed for lighter skin tones. The Fitzpatrick scale relies on subjective evaluations of hair and eye color alongside responses to ultraviolet radiation exposure, making it less reliable for darker skin tones [33,34]. In contrast, ITA° offers an objective and quantitative approach to skin color classification, making it more suitable for dermatological and cosmetic research. For instance, while we did not classify any volunteer as a Fitzpatrick skin type I, there is a Very Light ITA° subtype volunteer. The same is observed inversely; two volunteers have a Fitzpatrick skin type of VI, but our analysis has no Dark ITA° subtype volunteers.

In summary, our protocol demonstrates a clear correlation between the increase in white cast and the ZnO percentage in sunscreen formulations across different skin tones. These findings reinforce the objectivity of ITA° and support its continued use alongside CIEL*a*b* measurements for more reliable assessments [28]. The key observation is that white cast manifests differently depending on skin tone. This challenges the common assumption that only darker skin tones struggle with white cast; white cast manifests differently across skin tones but can be equally undesirable. The cosmetic industry must account for various skin tones when formulating mineral sunscreens. While some sunscreens provide reasonable white casts that are acceptable and aesthetically pleasing for certain skin tones, they often fail to achieve cosmetic elegance for others, leading to a critical gap in inclusivity within sunscreen development. Understanding and addressing the consumer’s concerns about cosmetic elegance is essential to ensure satisfactory and consistent use of sunscreens.

## Conclusion

In this study, we have developed a validated procedure for measuring and recording the white cast of mineral sunscreens containing ZnO in vivo and in vitro. This practical methodology, which involves objectively quantifying the white cast through colorimetry analysis and subjectively assessing it by photography and questionnaire, is designed to be easily applied in the development of mineral sunscreens. This procedure intends to optimize product development, allowing the early selection of formulations with a reduced white cast and enhanced cosmetic elegance for an improved consumer experience and, hopefully, user compliance.

## Material and methods

### Test material

Preparations containing various percentages of zinc oxide (ZnO) were used for the studies presented in this paper. Five test formulations containing 0.00%, 5.00%, 10.00%, 20.00%, or 30.00% ZnO were made in our laboratory (S4 Table). The ZnO used in these formulations is coated with triethoxycaprylylsilane, which has a particle size > 100 nm and was obtained from BASF (Florham Park, New Jersey, USA). The FDA allows up to 25% ZnO for over-the-counter sunscreen products for human use according to the Code of Federal Regulations: 21 CFR §352.10 (National Archives, 2024), but one of our formulations contained 30% ZnO. This was done according to the International Council for Harmonisation of Technical Requirements for Pharmaceuticals for Human Use (ICH) reportable ranges for common uses of analytical procedures, which states that for an assay of a drug substance or a finished (drug) product, the high-end of the reportable range is 120% of declared content or 120% of the upper specification limit [36]. The five test formulations were sent to contract laboratories for analytical assay of their ZnO percentage as determined by Inductively coupled plasma-optical emission spectroscopy (ICP-OES), and each formulation was confirmed accurate to the theoretical target. The test formulations were also sent for microbiological assay of total plate count, yeast and mold, coliforms, *E. coli*, *Pseudomonas* spp., *S. aureus*, *Salmonella* spp., and *C. albicans* (USP 61 and USP 62). There was no microbiological contamination present. Additionally, a primary dermal irritability 48-hour patch test study was conducted with 65 volunteers, and the test formulations did not elicit irritation. A commercially available Broad Spectrum SPF 30 with 14.7% ZnO and no other UV filter was used as the benchmark (BM) sample for comparison (S5 Table). For the in vivo and in vitro studies presented, each test formulation (a total of six) was applied at the clinically recommended dose of 2 mg/cm^2^.

### Colorimetry

The NIX Pro 2 Color Sensor (NixSensor, Hamilton, Ontario, Canada), along with the Nix Toolkit App available for iPhone, was the colorimeter used in this study for both in vitro and in vivo CIEL*a*b* measurements. The suitability of the NIX Pro 2 Color Sensor was checked daily before any experiment by conducting triplicate color measurements on black and white acrylic sheets (200 x 300 x 2 mm). The L*, a*, and b* relative standard deviations (RSD) were expected to be ≤ 2.0%.

### In vitro protocol for white cast measurement

Artificial skin (VITRO-SKIN) was cut into 6.0 x 6.0 cm square pieces and then hydrated according to the manufacturer’s instructions (IMS, Florida, USA) 16-24 hours before the experiment. Each test formulation was applied to a distinct piece of VITRO-SKIN. The VITRO-SKIN was inserted into a slide mount, and placed on top of a black acrylic sheet to produce a dark background before taking baseline color measurements. Three CIEL*a*b* measurements were taken per VITRO-SKIN at the top left, middle, and bottom right of the substrate (S3 Fig). Once the baseline measurements were taken, 2 mg/cm^2^ of the test samples were applied to their corresponding VITRO-SKIN. Post-application measurements were taken after a 15-minute rest to mimic usual sunscreen directions for human use. Once the sunscreens were set, three CIEL*a*b* measurements per sample were taken, similar to the baseline measurements. Moreover, the same procedure was performed with a white acrylic sheet as the background.

### In vivo protocol for white cast measurement

This protocol was a closed-label, double-blind study investigating how much white cast different ZnO formulations can cause in various skin types. Good Molecules, LLC, USA, sponsored and conducted this study between September and October 2024. Recruitment of potential volunteers was done between September 3^rd^, 2024 - October 1^st^, 2024, via email or text message by sending a flyer with a description of the study, basic requirements, and contact information to apply for the screening and subsequent participation in the study. The protocol complied with the guidelines of the Declaration of Helsinki and was approved by the Allendale Investigational Review Board (Old Lyme, CT, USA), which ensures that the study is conducted ethically and per regulatory requirements (Study Number GMCS#1-2024 approved on August 29th, 2024). All volunteers signed a written informed consent before initiating the study.

Healthy volunteers of legal age with a Fitzpatrick skin type between II and VI who were willing to comply with the procedures, methods, evaluations, and test product usage described in the written informed consent were included in the study. Volunteers were excluded if they had 1) dermatological conditions in the arms, such as Vitiligo, Rosacea, and Contact dermatitis, due to the potential interference with the study’s measurements; 2) a history of melanoma or treated skin cancer within the last five years due to the potential risk of skin damage; 3) tattoos, moles, or birthmarks covering the majority of their forearms due to the potential difficulty in measuring the white cast; 4) hairy forearms and an unwillingness to shave for the study due to the possible interference with the study’s measurements; and 5) a known allergy to any ingredient in the test formulations to ensure the safety of the volunteers and the accuracy of the study’s results.

Fourteen healthy adult volunteers started the study following the screening and recruitment process. However, one volunteer was excluded from the data analysis due to an ingredient allergy that prevented testing the benchmark sample and for not following the protocol. No other protocol deviations occurred during or after the study. The volunteers arrived at the study site with bare forearms, clean of skin care products, including sunscreen and moisturizers. Before measurements, the volunteers were given 15 minutes to acclimate to the ambient conditions. The research staff then cleaned the volunteers’ forearms with saline wipes, where they fixed six 3.0 x 3.0 cm silicone squares with hypoallergenic adhesive tape - one square per test formulation. Three baseline CIEL*a*b* measurements were taken at the top left, middle, and bottom right inside each silicone square, as per S3 Fig. The mean Individual Typology Angle (ITA°) was calculated from the CIEL*a*b* baseline measurements to categorize the volunteers’ skin color. The research staff then applied 2 mg/cm^2^ of each test formulation inside its respective silicone square (Fig 2A). After application, a 15-minute rest period was observed to let the formulations set on their skin before repeating CIEL*a*b* measurements.

Top-view photographs of the inner forearms were taken with an iPhone (X model) in a studio light box before and after the test samples application, as shown in Figs 2A and 2B. The before and after photographs comprised a color correction card (ColorChecker Classic by Calibrite LLC, Wilmington, Delaware, USA), which was used later to white balance the images with the aid of a photo-editing app (Snapseed by Google version 2.22, Mountain View, California, USA). During the study, the volunteers also completed a subjective questionnaire designed to collect their opinion on white cast and the test samples.

### Method Validation and Fit

The following set of analytical tests was conducted to validate our protocols. Linearity, accuracy, and precision for all CIEL*a*b* data were tested and calculated. For in vitro studies, we tested for both intra- and inter-day precision to confirm the repeatability and reliability of our measurements on different days with different staff. For in vivo studies, we tested only inter-day precision for the convenience of the study volunteers. We then conducted a comprehensive range of statistical tests, including the Shapiro-Wilk normality test, Pearson and Spearman Rho correlation, and Adjusted R-squared, to ensure the fit of our analysis. We used SPSS Version 30.0.0 for these statistics (IBM, Armonk, New York, USA). An 80-120% accuracy, a precision of < 10% RSD, and an adjusted R^2^ of > 0.650 were deemed acceptable for our studies.

### Statistical Analysis

The following set of descriptive statistics were provided for age: mean, SD, minimum, and maximum. Similarly, for gender, Fitzpatrick skin type, ethnicity and/or race, a detailed set of descriptive statistics is presented, along with their frequency expressed as number and/or percentage. Quantitative means and standard error of mean were calculated for the CIEL*a*b* data. In this study, we developed a white cast score to quantify the amount of white cast caused by sunscreens. For these white cast scores, the Zdifference (Zdiff) was calculated to determine whether correlation coefficients are equivalent or statistically different. First, we converted a pair of Pearson correlation coefficients into Fisher’s Z-scores to make them uniformly distributed. Then, we found the difference between these scores and adjusted for the variability in sample sizes to account for uncertainty. A larger Zdiff indicates a greater difference between the correlations when comparing this value to the critical threshold of Z = 1.96 (for 95% confidence). Apart from the statistical tests done for analytical validation, intraclass correlation coefficient (ICC) estimates and their 95% confident intervals were calculated using SPSS Version 30.0.0 (IBM, Armonk, New York, USA) based on an absolute-agreement, 2-way mixed-effects model. Graphs with mean, SEM, and two-way ANOVAs were done using PRISM 10 (GraphPad, Boston, Massachusetts, USA). We used R Studio Version 2024.09 (Posit PBC, Boston, Massachusetts, USA) and the RStudio package ggplot2, a powerful and widely used data visualization package in R, to generate the ITA° graph.

## Data Availability

Datasets related to this article can be found at doi: 10.17632/jfpzx7hcz2.1, hosted at Mendeley Data (Maldonado Lopez, Alexandra Gallagher, Emily Curry, Aiden Sahloff, Kylie da Silva Souza, Ivan (2025), ‘Data for: A Validated White Cast Scoring Method for Mineral Sunscreens’, Mendeley Data, V1, doi: 10.17632/jfpzx7hcz2.1).

https://doi.org/10.17632/jfpzx7hcz2.1

## Acknowledgments

The authors thank all volunteers for their participation in the study. We also thank the teams at Certified Laboratories and Kosmoscience for their support with microbiological, analytical, and clinical tolerance studies.

## Supporting Information

**S1 Fig. Individual Typology Angle Subtypes of Volunteers.** A total of thirteen (13) volunteers completed the study. The ITA°s range from Very Light to Brown subtypes.

**S2 Fig. In vitro Method of VITRO-SKIN over White Background is Less Sensitive to L* value Changes caused by Zinc Oxide Formulations.** (A) Only test formulations with ZnO percentages ranging from 10 - 30% show statistically significant changes in L* values after their application (p < 0.0001) n = 6. (B) Adjusted R^2^ was calculated to indicate the goodness of fit. VITRO-SKIN on white acrylic’s L* measurements (n = 30) ranged from 86 to 90. (C) Same graph as (b) but zoomed-in to observe L* value measurements clearly. BM = benchmark; ns = not significant. Error bars are shown as mean ± SEM.

**S3 Fig. Materials and methods for taking CIEL*a*b* measurements.** (A) Representative drawing of the in vitro model. The black rectangle is the black acrylic that serves as background. The gray square is the 6.0 x 6.0 cm VITRO-SKIN inserted into the slide mount (white outline). (B) Areas where CIEL*a*b* measurements were taken on VITRO-SKIN for the in vitro protocol and inside the silicone squares for the in vivo protocol. (C) Image of the NIX Pro 2 Color Sensor and App from the vendor’s website: https://www.nixsensor.com/product/nix-pro-color-sensor/.

**S1 Table. Inter-day Precision of In Vivo L* Measurements by Zinc Oxide Percentage.**

**S2 Table. Inter- and Intra-Day Precision of In Vitro L* Measurements by Zinc Oxide Percentage.**

**S3 Table. Demographics of the 13 Volunteers who Completed the In Vivo Protocol.**

**S4 Table. Laboratory Zinc Oxide Test Sample Formulations.**

**S5 Table. Commercially available Broad Spectrum SPF 30 with 14.7% ZnO.**

## Notes

### Competing Interest Statement

The study was fully sponsored by Good Molecules, LLC, a skincare brand. Both authors are employees of Good Molecules, LLC.

### Funding Statement

The author(s) received no specific funding for this work.

### Author Declarations

This study was performed in accordance with the Declaration of Helsinki. This human study was approved by Allendale Investigational Review Board (Old Lyme, CT, USA) - Study Number GMCS#1-2024 approved on August 29th, 2024. All adult participants provided written informed consent to participate in this study.

## References

1. Guan LL, Lim HW, Mohammad TF. Sunscreens and Photoaging: A Review of Current Literature. Am J Clin Dermatol. 2021 Nov;22(6):819–28.

2. Guy GP, Machlin SR, Ekwueme DU, Yabroff KR. Prevalence and Costs of Skin Cancer Treatment in the U.S., 2002−2006 and 2007−2011. Am J Prev Med. 2015 Feb;48(2):183–7.

3. Hughes MCB, Williams GM, Baker P, Green AC. Sunscreen and Prevention of Skin Aging: A Randomized Trial. Ann Intern Med. 2013 Jun 4;158(11):781.

4. Cole C, Shyr T, Ou-Yang H. Metal oxide sunscreens protect skin by absorption, not by reflection or scattering. Photodermatol Photoimmunol Photomed. 2016 Jan;32(1):5–10.

5. Serpone N. Sunscreens and their usefulness: have we made any progress in the last two decades? Photochem Photobiol Sci. 2021 Feb;20(2):189–244.

6. Ezekwe N, Pourang A, Lyons AB, Narla S, Atyam A, Zia S, et al. Evaluation of the protection of sunscreen products against long wavelength ultraviolet A1 and visible light-induced biological effects. Photodermatol Photoimmunol Photomed. 2024 Jan;40(1):e12937.

7. Addae AJ, Weiss PS. Standardizing the White Cast Potential of Sunscreens with Metal Oxide Ultraviolet Filters. Acc Mater Res. 2024 Apr 26;5(4):392–9.

8. Xiong M, Warshaw EM. Popular sunscreens marketed to individuals with skin of color: Cost, marketing claims, and allergenic ingredients. J Am Acad Dermatol. 2023 Apr;88(4):939–41.

9. Xu S, Kwa M, Agarwal A, Rademaker A, Kundu RV. Sunscreen Product Performance and Other Determinants of Consumer Preferences. JAMA Dermatol. 2016 Aug 1;152(8):920.

10. Akamine KL, Gustafson CJ, Davis SA, Levender MM, Feldman SR. Trends in Sunscreen Recommendation Among US Physicians. JAMA Dermatol. 2014 Jan 1;150(1):51.

11. Song H, Beckles A, Salian P, Porter ML. Sunscreen recommendations for patients with skin of color in the popular press and in the dermatology clinic. Int J Womens Dermatol. 2021 Mar;7(2):165–70.

12. Danovaro R, Bongiorni L, Corinaldesi C, Giovannelli D, Damiani E, Astolfi P, et al. Sunscreens Cause Coral Bleaching by Promoting Viral Infections. Environ Health Perspect. 2008 Apr;116(4):441–7.

13. Schneider SL, Lim HW. Review of environmental effects of oxybenzone and other sunscreen active ingredients. J Am Acad Dermatol. 2019 Jan;80(1):266–71.

14. Barone AN, Hayes CE, Kerr JJ, Lee RC, Flaherty DB. Acute toxicity testing of TiO2-based vs. oxybenzone-based sunscreens on clownfish (Amphiprion ocellaris). Environ Sci Pollut Res. 2019 May;26(14):14513–20.

15. Cocci P, Mosconi G, Palermo FA. Sunscreen active ingredients in loggerhead turtles (Caretta caretta) and their relation to molecular markers of inflammation, oxidative stress and hormonal activity in wild populations. Mar Pollut Bull. 2020 Apr;153:111012.

16. Levine A. Reducing the prevalence of chemical UV filters from sunscreen in aquatic environments: Regulatory, public awareness, and other considerations. Integr Environ Assess Manag. 2021 Sep;17(5):982–8.

17. Battistin M, Pascalicchio P, Tabaro B, Hasa D, Bonetto A, Manfredini S, et al. A Safe-by- Design Approach to “Reef Safe” Sunscreens Based on ZnO and Organic UV Filters. Antioxidants. 2022 Nov 9;11(11):2209.

18. Luckiesh M. Protective skin coating for the prevention of sunburn. J Am Med Assoc. 1946 Jan 5;130(1):1.

19. Smijs T, Pavel. Titanium dioxide and zinc oxide nanoparticles in sunscreens: focus on their safety and effectiveness. Nanotechnol Sci Appl. 2011 Oct;95.

20. Singh P, Nanda A. Enhanced sun protection of nano-sized metal oxide particles over conventional metal oxide particles: An *in vitro* comparative study. Int J Cosmet Sci. 2014 Jun;36(3):273–83.

21. Mitchnick MA, Fairhurst D, Pinnell SR. Microfine zinc oxide (Z-Cote) as a photostable UVA/UVB sunblock agent. J Am Acad Dermatol. 1999 Jan;40(1):85–90.

22. Schneider SL, Lim HW. A review of inorganic UV filters zinc oxide and titanium dioxide. Photodermatol Photoimmunol Photomed. 2019 Nov;35(6):442–6.

23. Xu L, Zhou Y, Amin S. Chapter 13. Polymer Colloids for Cosmetics and Personal Care. In: Priestley R, Prud’homme R, editors. Soft Matter Series. Cambridge: Royal Society of Chemistry; 2019. p. 399–417. Available from: https://books.rsc.org/books/monograph/800/chapter-abstract/537275/

24. Barnard AS. One-to-one comparison of sunscreen efficacy, aesthetics and potential nanotoxicity. Nat Nanotechnol. 2010 Apr;5(4):271–4.

25. Ly BCK, Dyer EB, Feig JL, Chien AL, Del Bino S. Research Techniques Made Simple: Cutaneous Colorimetry: A Reliable Technique for Objective Skin Color Measurement. J Invest Dermatol. 2020 Jan;140(1):3–12.e1.

26. Richer V, Kharazmi P, Lee TK, Kalia S, Lui H. Quantifying the visual appearance of sunscreens applied to the skin using indirect computer image colorimetry. Photodermatol Photoimmunol Photomed. 2018 Mar;34(2):130–6.

27. Hasnul Hadi MH, Ker PJ, Thiviyanathan VA, Tang SGH, Leong YS, Lee HJ, et al. The Amber-Colored Liquid: A Review on the Color Standards, Methods of Detection, Issues and Recommendations. Sensors. 2021 Oct 16;21(20):6866.

28. Del Bino S, Bernerd F. Variations in skin colour and the biological consequences of ultraviolet radiation exposure. Br J Dermatol. 2013 Oct;169:33–40.

29. Everett JS, Budescu M, Sommers MS. Making Sense of Skin Color in Clinical Care. Clin Nurs Res. 2012 Nov;21(4):495–516.

30. Del Bino S, Ito S, Sok J, Nakanishi Y, Bastien P, Wakamatsu K, et al. Chemical analysis of constitutive pigmentation of human epidermis reveals constant eumelanin to pheomelanin ratio. Pigment Cell Melanoma Res. 2015 Nov;28(6):707–17.

31. Wu Y, Tanaka T, Akimoto M. Utilization of Individual Typology Angle (ITA) and Hue Angle in the Measurement of Skin Color on Images. Bioimaging Society; 2020. Available from: 10.11169/bioimages.28.1

32. Ibraheem NA, Hasan MM, Khan RZ, Mishra PK. Understanding color models: a review. ARPN Journal of science and technology. 2012;2(3):265–75.

33. Fitzpatrick TB. The validity and practicality of sun-reactive skin types I through VI. Arch Dermatol. 1988 Jun;124(6):869–71.

34. Ware OR, Dawson JE, Shinohara MM, Taylor SC. Racial limitations of fitzpatrick skin type. Cutis. 2020 Feb;105(2):77–80.

35. National Archives. 21 CFR 352.10 Sunscreen active ingredients. In: Code of Feredral Regulations. Available from: https://www.ecfr.gov/current/title-21/chapter-I/subchapter-D/part-352/subpart-B/section-352.10

36. 36. US FDA. Q2(R2) Validation of Analytical Procedures Guidance for Industry. Available from: https://www.fda.gov/regulatory-information/search-fda-guidance-documents/q2r2-validation-analytical-procedures

